# Age and sex disaggregation of crude excess deaths during the 2020 spring COVID-19 outbreak in Spain with an estimate of infection fatality rate

**DOI:** 10.1101/2020.08.06.20169326

**Authors:** José María Martiín-Olalla

**Affiliations:** Universidad de Sevilla. Facultad de Física. Departamento de Física de la Materia Condensada. ES41012 Sevilla. Spain

**Keywords:** mortality excess, z-score, EuroMoMo, INE, Instituto de Salud Carlos III

## Abstract

Spanish official records of mortality and population during the 21st century are analyzed to determine the age-sex specific crude death rate in the 2020 spring (week 10 to week 21) COVID-19 outbreak.

Age-sex specific cumulative death rates can be modelled by a Poission regression with rate linearly variying with calendar year from which age-sex specific reference value for 2020 are obtained.

Excess death rate increases exponentially with age showing a doubling time [4.1, 4.9]a (female) and [4.8, 5.4]a (male). Age specific infection fatality rate doubling times below age 70 years are reported as [4.7, 8.8]a (female) and [4.8, 6.6]a (male).

Infection fatality rate for people aged more than 80 is discussed in relation to the shares of people living in institutionalized long term care facilities.

## 1. INTRODUCTION

The illness designated COVID-19 caused by the severe acute respiratory syndrome coronavirus 2 (SARS-CoV-2) caught worldwide attention since its identification in late 2019[1]. Spain is one of the European countries most impacted by the disease during the spring of 2020[2]. Confirmed COVID-19 cases climbed up to several hundred thousands —some few thousands per one million population— and confirmed COVID-19 deaths to some 28000 or six hundred deaths per one million population.[3] Lock-down measures came into effect on March 16 to help diminishing the outbreak.

Total excess deaths or crude excess deaths is a key quantity to understand the acute impact of a pandemic[4]. Its determination sensibly requires a reference mortality or baseline and a time interval over which the excess is determined. Baseline is determined from previous records of mortality. European National Statistical Institutes and Eurostat are doing a great effort in disseminating weekly deaths in Europe in the past years.

Age specific and sex specific analyses are of great interest since they addressed the question to whom the diseases impact the most[5]. Age specific analysis of mortality in Spain is heavily influenced by the strong impact of the disease within nursing homes[6]. This manuscript takes official records of weekly crude deaths in Spain during the 21st century and population records to ascertain the impact of the COVID-19 in age-sex specific death rates, including a discussion on age specific infection fatality rates.

## 2. DATA SETS

In April 2020 Eurostat set up “an exceptional temporary data collection on total week deaths in order to support the policy and research efforts related to COVID-19”[7]. The data collection is disaggregated by sex, five year age group and NUTS regions in several countries of the European Union and elsewhere. Data are provided by National Statistical Institutes on a voluntary basis. The 2020 weekly deaths still have the flag of “estimate”.

Age group population values until January 1, 2019 for Spain were also collected from Eurostat demo_pjan table. This table disaggregates age year by year which allows to build up five year age groups matching to the weekly death data. Linear interpolation was used to compute population a weekly basis.

Total population values for January 1,2020 can be collected from table ts00001 in Eurostat. Unluckily there is no age disaggregation. However these figure were obtained in Spain from the table 31304 at the Instituto Nacional de Estadística. For the Dutch set the last two available shares of population for every age group were used to extrapolate the shares of population in 2020.

Eurostat requested the National Statistical Institutes the transmission of “a back series weekly deaths for as many years as possible, recommending as starting point the year 2000”. In this manuscript Spanish and Dutch weekly deaths will be analyzed from 2001 on-wards. As Eurostat points out “a long enough time series is necessary for temporal comparisons and statistical modelling”.

## 3. RESULTS

In specific death rate analysis the population *N* is assumed to be a blending, a mixture, or a multicomponent system composed of several age groups and sex groups of size *N_i_* whose behaviour in relation to death is homogeneous under some circumstances. Age-sex specific weekly death rate[8] *d_i_* is nothing but the ratio between weekly deaths *D_i_* within a group *i* and its population *N_i_*. All age-sex specific weekly deaths sum up the total weekly deaths 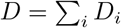. Also considering the definition of age-sex specific death rate this can be written as 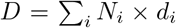 from which the total weekly death rate *d* = *D/N* can be obtained as 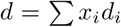 where *x_i_* = *N_i_/N* is the shares of population by age-sex group. Total weekly death rate is then the weighted average of age-sex specific death rates.

Results can also be partially grouped, ie. death rates for both male and female yields the total sex category.

Figure 1 shows the male (top) and female (bottom) age specific weekly crude death rate *d_i_* in Spain for 19 age groups (color from a gradient palette) the total —age unspecific— weekly crude death rate *d* (orange). Death rates are distributed exponentially following the Gompertz law[9] *d(a)* ∝ 2*^a/τ^* where *a* is the age and *τ* is the doubling time. This empirical law suggests the predominance of age specific contributions to the cause of death instead of external causes like wars, murders, plagues or the like.

**FIG. 1.**
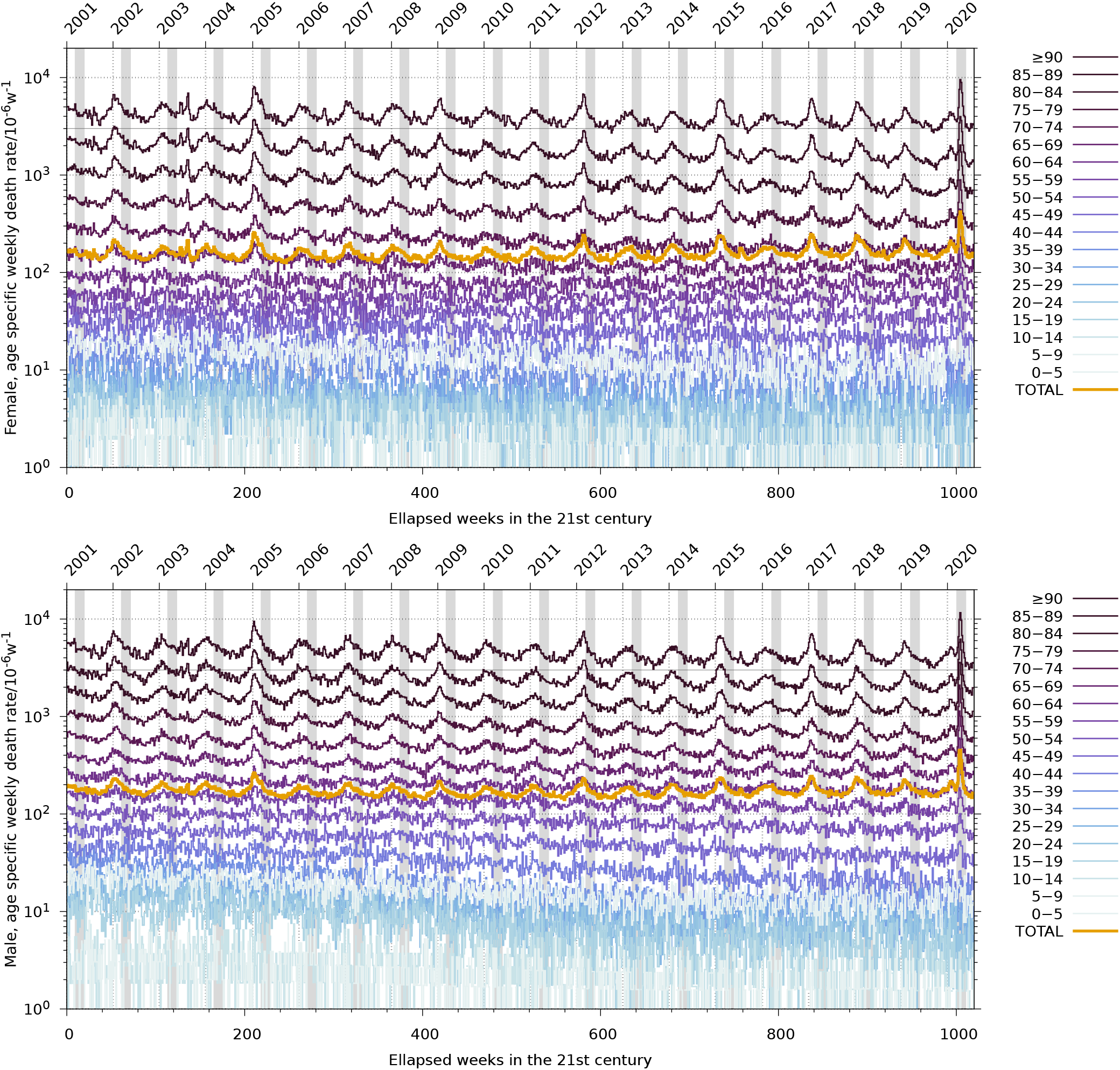
Age specific, sex specific —female (top) and male (bottom)—, weekly crude death rate in Spain during the 21st century in a gradient color. Orange line shows the total weekly crude death rate. Gray strip bands locate W10 to W21.

Following the ideas in Ref. [10] this work will analyze the cumulative weekly death rate from W10 to W21 in the 21st century. This interval is noted by strip bands in Figure 1. In 2020 W10 to W21 extends from Monday March 2, 2020 to Sunday May 24, 2020. In this period of time weekly deaths exhibited an extremely large anomaly in Spain (*z*– score equal to 16). The data collection allows to compute the cumulative deaths recorded in this period for the past years and for every age group and sex. Scaling by the group population at the given year gives us the cumulative death rate in this period and year.

Figure 2 shows the results of this statistics in Spain. Every panel shows age specific male (blue) and female (orange) death rates against calendar year. Vertical axis is normalized by removing the both sex (total) average death rate and by scaling by the total standard deviation. Black lines the central tendency of a full model Poisson regression for data 2001 to 2019 (*n* = 19) with a response linearly depending on calendar year. The tendency is extended until 2020 to give the predicted reference value from the model in that year.

**FIG. 2.**
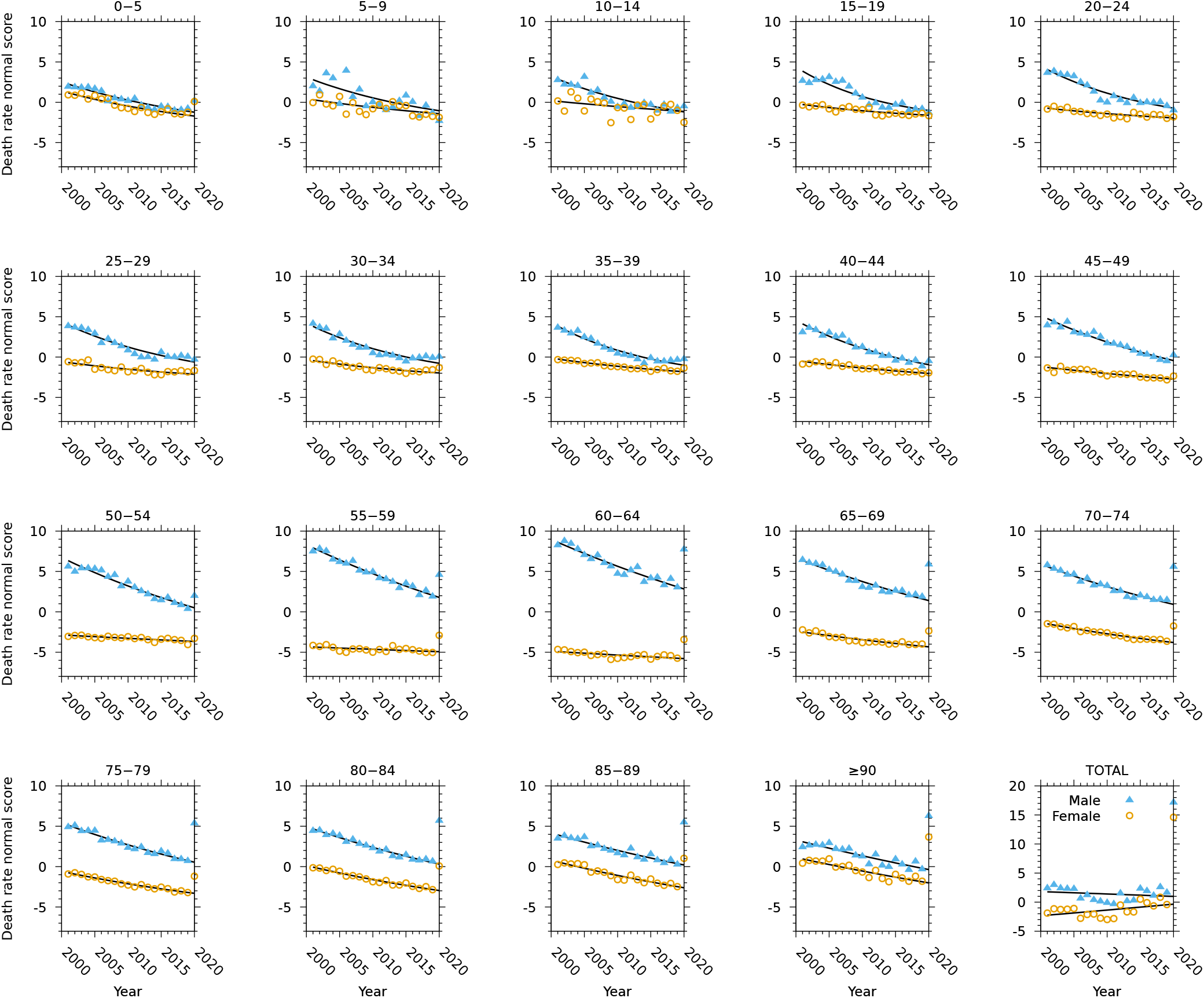
Age and sex specific death rates from W10 to W21 in Spain. For every age group male death rate is noted in blue (triangles); female in orange (circles). Black line shows the central tendency from a Poisson regression that fitted death rates and calendar year from 2001 to 2019. The predicted value in 2020 is the reference for the model. Notice 2020 observed values lying outside the prediction interval in the two bottom-most rows. Bottom-right most panel displays the total age group death rates. The normal score was obtained removing the total sex sample average value 2001-2019 and scaling by the total sex standard deviation.

It is worthy to note that the youngest age group exhibits an anomalously high increase in 2020 (see also Table 1 in Ref. [6]). However a close inspection of the signal in Spain only reveals a sudden upraise in mid 2019 after which the signal fluctuates normally, including W10 to W21 in 2020. As an hypothesis it might be possible that fake 0 age deaths are initially reported and attributed to this age group. Later they are cleansed by the National Statistics Institute after close inspection of records. Data after mid 2019 still lacks this upgrade.

**TABLE 1.**
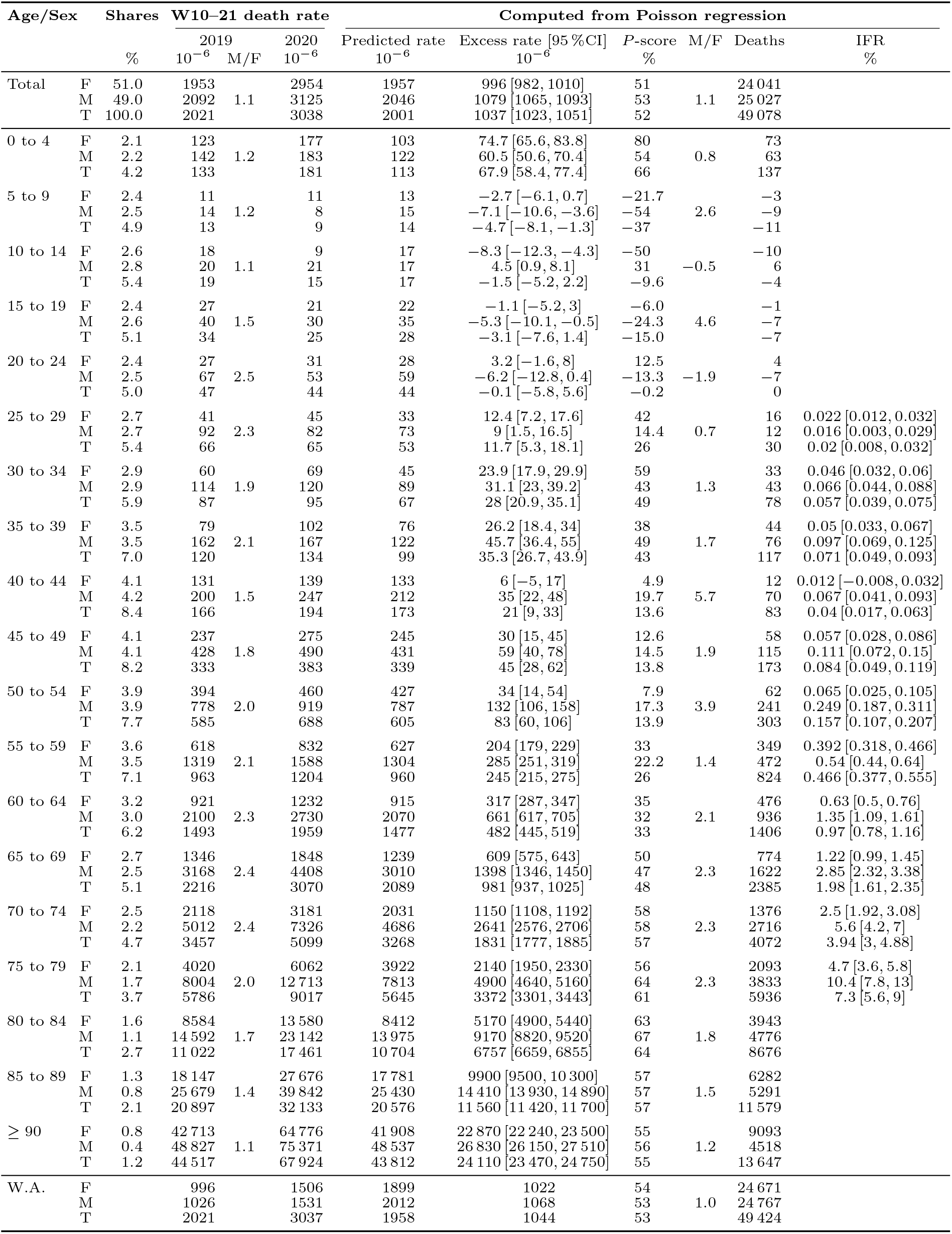
Age specific cumulative death rate by sex (F, female; M, male; T, total) in Spain from W10 to W21. First column shows the age-sex specific shares of population *x* in 2020 when *N* = 47 329 981. Then the observed cumulative death rate in 2019 and male to female ratio, folowed by the observed cumulative death rate in 2020. Then results from a full model linear Poisson regression (see Figure 2): the predicted value *r* and the excess *e* = *o* − *r* followed by 95 % confidence interval, *P*–score *e/r*, and the male to female ratio. Then age-sex specific total deaths *e* × *x* × *N* and the infection fatality rate IFR as the ratio of excess death rate to the shares of seroprevalence from Ref. [6] First three rows report overall age unspecific excesses. Last three rows report weight averages and grand totals of age specific excesses. Both differ in 1 %.

Table 1 summarizes the results displayed in Figure 2. It first lists the shares of population in 2020. Then the observed cumulative death rate *o* in 2019 followed by the male to female death rate ratio. Then the observed cumulative death rate in 2020. Next group of columns displays the results in Figure 2: first the predicted value or reference *r* for 2020 followed by the death rate excess *e* = *o* — *r* and the *P*-score *e/r*. Then the male to female ratio. With few exceptions in the middle ages 2020 excess death rate male to female ratio and 2019 death rate male to female ratio are roughly coincidental.

This is followed by excess age-sex excess deaths *e* × *x* × *N* and in the final column data by the age-sex specific infection fatality rate IFR. Values for IFR are computed as the ratio of excess death rate to the shares of IgG+ seroprevalence[6, 11] with confidence intervals given by the delta method. Aged groups of population are not considered because seroprevalence is only representative of non-institutionalized population (see Refs [6, 11]) and large shares of aged population live in institutionalized long-term care facilities (see also Section 4.2).

There are two totals in Table 1. First three rows report results for the age unspecific total group. This is the analysis of the sum of age specific deaths and it is equivalent to that reported in Ref. [10] except for the fact that there excess were taken relative to the average death rate in 2001-2019. It should be noted that age-unspecific male cumulative death rate sustains the null hypothesis of no relationship with calendar year (*p >* 0.4), whereas the age- unspecific female cumulative death rate does not (*p* = 0.02). However, to keep consistency with age-specific analysis this work considers a full linear model in every Poisson regression. This brings age unspecific Spanish excess deaths from ~ 50 000[10] to ~ 49 000 (Table 1).

Last three rows report the weight average results, which is the weighted sum of excess for age specific analysis. Both totals differ in 1 %.

## 4. DISCUSSION

### 4.1. Doubling times

Mimicking Figure 2 one may think in plotting age-sex specific cumulative death rates as a function of age group for every year in the collection. As an example first two columns in Table 1 show a steadily progressive increase with age group for every sex. Same happens for excess death rates and for IFR in Table 1.

Figure 3 shows these tendencies in a semilogarithmic plot. Every panel shows exponential increase (Gompertz law) with doubling time slightly larger than 6 (death rates) and 5 (excess death rates). Since seroprevalence[11] only changes smoothly with age, IFR replicates the logarithmic behaviour of age specific excess deaths.

**FIG. 3.**
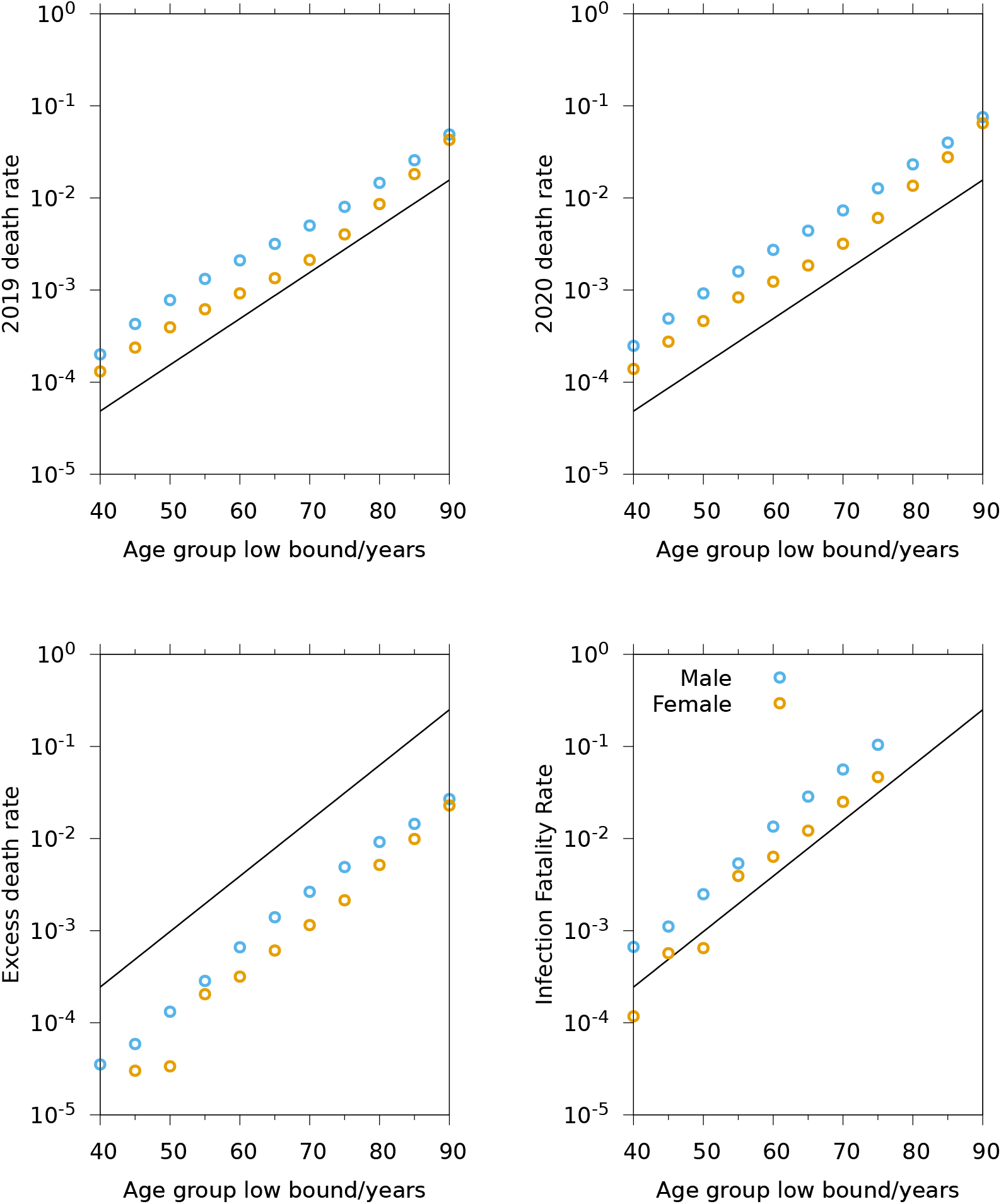
Semilogarithmic plot of the observed age specific death rates in 2019 and 2020; excess death rate in Spain. Blue circles shows male statistics; orange circles, female statistics. Straight line shows doubling times *τ* = 6 a (death rates) and *τ* = 5 a (excess death rate). Excess death rate confidence intervals shown in Table 1 are close to point size in this figure. They are not printed for the sake of clarity. IFR confidence intervals are exceedingly large except for the last four data points due to the logarithmic axis. Doubling times are reported in Table 2.

Table 2 summarizes the results of the exponential fitting *y* ∝ 2*^a/τ^*, where *a* is the low bound of age group and *y* is any of the responses. A doubling rate close to 5 a means that excess death rates doubles in every step of increase in the age group staircase from 40-45 on and it translates into a log-linear slope equal to ~ log(2)/(5a) ~ 0.126 a^-1^, in accordance with Figure 3 in Ref. [5].

**TABLE 2.**
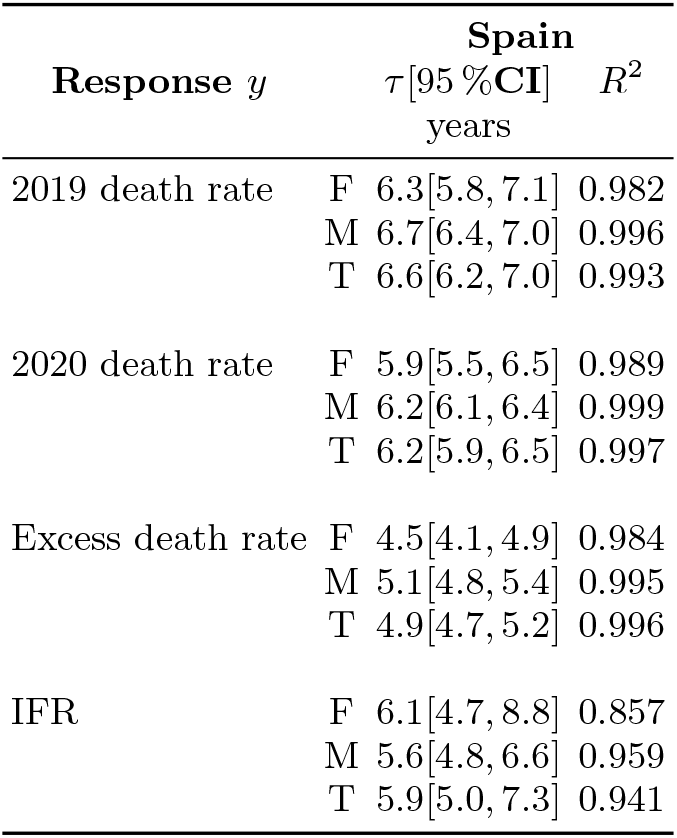
Doubling times *y* ∝ 2*^a/τ^* —where *a* is age group low bound— of death rates, excess death rate and IFR and Pearson’s correlation coefficient in Spain and Netherlands. Only population aged 40 or more enters in this analysis (*n* = 11), see Figure 3.

### 4.2. IFR in aged groups of population

Following the ideas by Pastor-Barriuso *et al*. [6] the ratio of all-cause excess deaths to seroprevalence is not representative of the IFR in the most aged groups of population due to shares of population living in institutionalized long-term care facilities which did not enter in the seroprevalence study.

In order to give some context Abellán García *et al*. [12] estimated 322 000 persons living in Spanish long term care facilities. It is only a 3% of Spanish population aged more than 65 years. However, *N*_1_ = 19 681 deaths[6] related to COVID-19 were reported in institutionalized long-term care facilities, which makes 42 % of the 46 295 excess deaths in population aged more than 65 years (see Table 1).

Pastor-Barriuso *et al*. [6] discussed in detail this issue. They considered *N*_2_ = 44459 excess death as reported from the Monitor of Mortality[13]) from which they excluded *N*_1_ to give an estimate of excess deaths in Spanish non-institutionalized population. IFR was then computed as *d_n_/s_n_* where *s_n_* is the shares of seroprevalence within non-institutionalized population and *d_n_* is the excess death rate in non-institutionalized population.

Infection fatality rate reported in Ref. [6] (Table 1) is listed in the first column data of Table 3. The original IFR can be transformed into *d_n_/(s_n_* + *d_n_*). This metric is relevant if *s_n_* ~ *d_n_* as it happens in the most aged groups when *d* climbs up to few percent.

**TABLE 3.** Age and sex specific IFR in Spain from Pastor-Barriuso *et al*. [6] and from this work. First column data lists the shares of population living in long-term care facilities *x*. Original data in Ref. [6] are *d_n_/s_n_* where *d_n_* is age-sex specific death rate in non-institutionalized population (overall deaths adds up to 24 778) and *S_n_* is the shares of seroprevalence in noninstitutionalized population. Original data are reformed into *d_n_* / (*s_n_* + *d_n_*). Reformed data are augmented into 1. 2 × *d_n_* / (*s_n_* +*d_n_*). This work IFR displays *d*/((1 − *x*) · *s_n_* + *d*) where *d* is excess death rate. Overall non-institutionalized excess deaths adds up to 29 743, which differs in 20 % from the value used by Pastor-Barriuso *et al*. and justifies the augmented value.

| Age | Inst Pop[12, 14]<br>$x$<br>% | IFR Pastor-Barriuso *et al.* [6] | | | This work | | |
| --- | --- | --- | --- | --- | --- | --- | --- |
|  |  | Original<br>% | Reformed<br>% | Augmented<br>% | IFR<br>% | Excess rate<br>% | Deaths |
| Male |  |  |  |  |  |  |  |
| 0–9 | 0.0 | 0.0 | 0.0 | 0.0 | 0.1 | 0.0 | 54 |
| 10–19 | 0.0 | 0.0 | 0.0 | 0.0 | 0.0 | 0.0 | –1 |
| 20–29 | 0.0 | 0.0 | 0.0 | 0.0 | 0.0 | 0.0 | 4 |
| 30–39 | 0.0 | 0.0 | 0.0 | 0.0 | 0.1 | 0.0 | 119 |
| 40–49 | 0.0 | 0.1 | 0.1 | 0.1 | 0.1 | 0.0 | 185 |
| 50–59 | 0.0 | 0.3 | 0.3 | 0.4 | 0.4 | 0.0 | 713 |
| 60–69 | 0.2 | 1.6 | 1.6 | 1.9 | 2.0 | 0.1 | 2558 |
| 70–79 | 1.4 | 6.1 | 5.8 | 6.9 | 7.2 | 0.4 | 6549 |
| ≥ 80 | 8.9 | 16.4 | 14.1 | 16.9 | 24.8 | 1.4 | 14 585 |
| Female |  |  |  |  |  |  |  |
| 0–9 | 0.0 | 0.0 | 0.0 | 0.0 | 0.1 | 0.0 | 70 |
| 10–19 | 0.0 | 0.0 | 0.0 | 0.0 | 0.0 | 0.0 | –12 |
| 20–29 | 0.0 | 0.0 | 0.0 | 0.0 | 0.0 | 0.0 | 19 |
| 30–39 | 0.0 | 0.1 | 0.0 | 0.1 | 0.0 | 0.0 | 77 |
| 40–49 | 0.0 | 0.0 | 0.0 | 0.0 | 0.0 | 0.0 | 70 |
| 50–59 | 0.0 | 0.2 | 0.2 | 0.2 | 0.2 | 0.0 | 411 |
| 60–69 | 0.2 | 0.6 | 0.6 | 0.7 | 0.9 | 0.0 | 1250 |
| 70–79 | 1.4 | 2.7 | 2.6 | 3.1 | 3.4 | 0.2 | 3469 |
| ≥ 80 | 8.9 | 6.5 | 6.1 | 7.3 | 19.1 | 1.1 | 19 317 |

Pastor-Barriuso *et al*. started with an overall excess death in non-institutionalized population equal to *D*_1_ = *N*_2_ *− N_1_* = 24 778. In this work age-sex specific excess deaths adds up 49424, which makes *D*_2_ = 49 424 − 19 681 = 29 743 overall excess deaths in non-institutionalized population. They differ in *D*_2_/*D*_1_ = 1.20. As a result IFR in Ref. [6] must be augmented in a factor 1.20 to be compared.[15]

On the other hand overall IFR can be computed as the ratio of excess deaths (see Table 1) to seroprevalance reported in Ref. [6]. This seroprevalence must be corrected to account for the shares of institutionalized population which did not enter in the seroprevalence study. IFR can then be expressed as *d*/(*d* + *s_n_*(1 − *x*)). The share *x* can be obtained from the estimates for Spanish population living in long-term care facilities[12, 14]. The (1 − *x*) accounts for the fact that *d* is scaled by resident population while *s_n_* is scaled by non-institutionalized population.

Taking into account all this, Table 3 shows that the only relevant difference is observed for the most aged group. On the first hand this group exhibits 16.9% (male) and 7.3% (female) for non-institutionalized IFR, and on the other hand, it makes 24.8% (male) and 19.1 % (female) for overall IFR. Understandably the issue arises only if the shares of population living in long-term care facilities is large enough. The difference is huge —a factor ×2.6— for female population which highlights the larger shares of population at these ages and the larger shares of people living in nursing homes.

IFR reported in this work is larger because it still lacks the shares of infected survivors living in long-term care facilities. To the best of my knowledge this is yet an unknown quantity and it is the central point of Ref. [6]. In other words, IFR reported in Table 3 would be accurate only if every infection in long-term care facilities had ended in a death, which is not the case.

More sensibly the IFR could be computed as *d*/((1 − *x*) · *s_n_* + *d* + *y*), where *y* is the (unknown) ratio of infected survivors within institutionalized population to the overall population in the group. Figure 4 shows this function for male and female population. The figure highlights the striking difference in the female group: not even when every surviving woman aged ≥ 80 living in long-term care facilities were infected, the overall IFR would fall to the value reported for non-institutionalized women. This suggests an specific impact of COVID-19 in institutionalized females that goes beyond of the size of the infection.

**FIG. 4.**
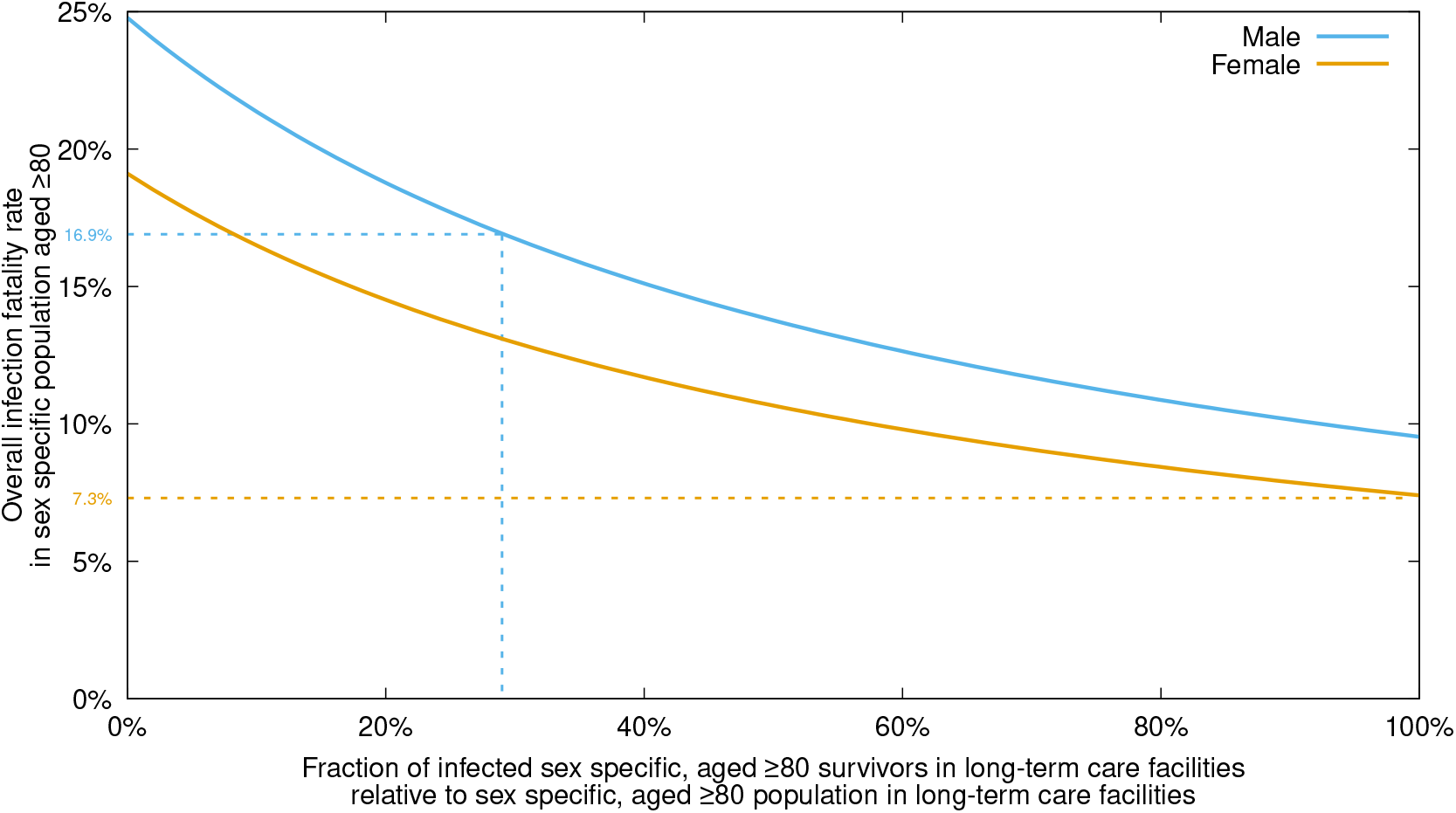
Overall infection fatality rate in Spain during the spring COVID-19 outbreak for male (blue) and female (orange) population aged ≥ 80 as a function of the fraction of (unknown) infected survivors living in long-term care facilities relative to population living in long-term care facilities. Broken lines show the values reported by Pastor-Barriuso *et al*. [6] for sex specific population aged ≥ 80 not living in long-term care facilities.

On the other, if 30 % of male institutionalized population aged ≥ 80 were infected survivors then the overall IFR would fall to the value reported for non-institutionalized population. Arguably this might be a highest bound for the shares of male infected survivors since, otherwise, overall IFR would be smaller than IFR in non-institutionalized population.

Alternatively, IFR can be computed for non-institutionalized population aged more than 80 years. From Table 3 there are *N*_3_ = 33 902 all-sex excess death in this group of population. The overwhelming majority of *N*_1_ must be assigned to this group of population because: (1) death rates increase exponentially as age increases and (2) the shares of population living in institutionalized facilities increase with age. Therefore a rough estimate of non-institutionalized excess death for people aged more than 80 years is *N*_4_ = *N*_3_ − *N*_1_ × 0.95 = 14 200 or an excess death rate equal to d_4_ = 5800 × 10^-6^. Considering the seroprevalence of this age group the infection fatality rate for non-institutionalized,

## 5. CONCLUSIONS

Spanish age-sex specific cumulative death rate in the period W10 to W21 during the 21st century can be modelled by a Poisson regression with a rate linearly depending on calendar year. Age-sex specific excess death rates in Spain during the spring 2020 (W10 to W21) COVID-19 outbreak follows Gompertz exponential law with doubling times in the range [4.1, 4.9]a (female) and [4.8, 5.4]a (male). Age specific infection fatality rate doubling times below age 70 years are reported as [4.7, 8.8]a (female) and [4.8, 6.6]a (male) in accordance with previous reports[5].

An upper bound for the over-all infection fatality rate in population aged more than 80 is reported as 25 % (male) and 19% (female). For non-institutionalized population IFR is reported as 12% (sex unspecific).

## Data Availability

All data come from public repositories at Eurostat or National Statistical Institutes.

## 7. DATA AVAILABILITY

Every piece of data in this work comes from public collection found in Eurostat and the Instituto Nacional de Estadística.

## ACKNOWLEDGMENTS

The palette used in Figure 1 is named “dense”, from cmocean palettes by Kristen Thyng https://matplotlib.org/cmocean/.

The author thanks National Statistics Institutes and Eurostat for releasing public records of weekly deaths. I knew of Eurostat effort from a tweet posted by Kiko Llaneras https://twitter.com/kikollan/status/1288830168925188096 on July 30, 2020.

This work was not founded.

This work benefited from free software running on xubuntu 18.04.1LTS. Data bases were imported into GNU octave-4.2.2 (https://www.gnu.org/software/octave/). Poisson regressions were performed by R-3.6.3 (https://www.r-project.org/). Pictures were developed thanks to gnuplot-5.2.2 (http://www.gnuplot.info/). The manuscript was typeset in GNU emacs-25.2.2 (https://www.gnu.org/software/emacs/) assisted by AUCTEX(https://www.gnu.org/software/auctex/). Data in pictures and in tables have been exported directly from octave.

This project started on July 31, 2020.

## REFERENCES

1 N. Zhu, D. Zhang, W. Wang, X. Li, B. Yang, J. Song, X. Zhao, B. Huang, W. Shi, R. Lu, P. Niu, F. Zhan, X. Ma, D. Wang, W. Xu, G. Wu, G. F. Gao, and W. Tan, New England Journal of Medicine 382, 727 (2020).

2 A. García-Basteiro, C. Alvarez-Dardet, A. Arenas, R. Bengoa, C. Borrell, M. Del Val, M. Franco, M. Gea-Sanchez, J. J. G. Otero, B. G. L. Valcárcel, I. Hernández, J. C. March, J. M. Martin-Moreno, C. Menendez, S. Minue, C. Muntaner, M. Porta, D. Prieto-Alhambra, C. Vives-Cases, and H. Legido-Quigley, The Lancet 0 (2020), 10.1016/S0140-6736(20)31713-X.

3 Instituto de Salud Carlos III COVID-19 in Spain https://cnecovid.isciii.es/covid19 (accessed July 2020).

4 J. Aaron, J. Mullbauer, C. Giattino, and H. Ritchie, “A pandemic primer on excess mortality statistics and their comparability across countries,” (2020).

5 A. T. Levin, K. B. Cochran, and S. P. Walsh, medRxiv, 2020.07.23.20160895 (2020).

6 R. Pastor-Barriuso, B. Pérez-Gómez, M. A. Hernán, M. Pérez-Olmeda, R. Yotti, J. Oteo, J. L. Sanmartín, I. Leán- Gomez, A. Fernández-García, P. Fernández-Navarro, I. Cruz, M. Martín, C. Delgado-Sanz, N. Fernández De Larrea, J. León Paniagua, J. F. Muñoz-Montalvo, F. Blanco, A. Larrauri, and M. Pollán, medRxiv, 2020.08.06.20169722 (2020).

7 https://ec.europa.eu/eurostat/cache/metadata/en/demomwk_esms.htm.

8 This manuscript will print death rates as a number times 10^-6^ where the number stands for the mortality per one million population.

9 B. Gompertz, Philosophical Transactions of the Royal Society of London 115, 513 (1825).

10 J. M. Martín-Olalla, medRxiv, 2020.07.22.20159707 (2020).

11 M. Pollán, B. Pérez-Gómez, R. Pastor-Barriuso, J. Oteo, M. A. Hernán, M. Pérez-Olmeda, J. L. Sanmartín, A. Fernández-García, I. Cruz, N. Fernández de Larrea, M. Molina, F. Rodríguez-Cabrera, M. Martín, P. Merino-Amador, J. León Paniagua, J. F. Muñoz-Montalvo, F. Blanco, R. Yotti, F. Blanco, R. Gutierrez Fernandez, M. Martín, S. Mezcua Navarro, M. Molina, J. F. Muñoz-Montalvo, M. Salinero Hernández, J. L. Sanmartín, M. Cuenca-Estrella, R. Yotti, J. León Paniagua, N. Fernández de Larrea, P. Fernández-Navarro, R. Pastor-Barriuso, B. Pérez-Gómez, M. Pollán, A. Avellón, G. Fedele, A. Fernández-García, J. Oteo Iglesias, M. T. Pérez Olmeda, I. Cruz, M. E. Fernandez Martinez, F. D. Rodríguez-Cabrera, M. A. Hernán, S. Padrones Fernández, J. M. Rumbao Aguirre, J. M. Navarro Marí, B. Palop Borrvs, A. B. Pérez Jiménez, M. Rodríguez-Iglesias, A. M. Calvo Gascon, M. L. Lou Alcaine, I. Donate Suárez, O. Suárez Alvarez, M. Rodríguez Pérez, M. Cases Sanchís, C. J. Villafafila Gomila, L. Carbo Saladrigas, A. Hurtado Ferníndez, A. Oliver, E. Castro Feliciano, M. N. González Quintana, J. M. Barrasa Fernández, M. A. Hernández Betancor, M. Hernández Febles, L. Martín Martín, L.-M. López López, T. Ugarte Miota, I. De Benito Población, M. S. Celada Pérez, M. N. Vallés Fernández, T. Maté Enriquez, M. Villa Arranz, M. Domínguez-Gil González, I. Fernández-Natal, G. Megías Lobón, J. L. Muñoz Bellido, P. Ciruela, A. Mas i Casals, M. Doladé Botiías, M. A. Marcos Maeso, D. Campo, A. Félix de Castro, R. Limón Ramírez, M. F. Elías Retamosa, M. Rubio González, M. S. Blanco Lobeiras, A. Fuentes Losada, A. Aguilera, G. Bou, Y. Caro, N. Marauri, L. M. Soria Blanco, I. del Cura González, M. Hernández Pascual, R. Alonso Fernández, P. Merino-Amador, N. Cabrera Castro, A. Tomás Lizcano, C. Ramírez Almagro, M. Segovia Hernández, N. Ascunce Elizaga, M. Ederra Sanz, C. Ezpeleta Baquedano, A. Bustinduy Bascaran, S. Iglesias Tamayo, L. Elorduy Otazua, R. Benarroch Benarroch, J. Lopera Flores, and A. Vazquez de la Villa, The Lancet 0 (2020), 10.1016/S0140-6736(20)31483-5.

12 A. Abellán García, P. Aceituno Nieto, I. Fernández Morales, D. Ramiro Fariñas, and R. Pujol Rodríguez, “Una estimación de la población que vive en residencias de mayores,” (2020).

13 See https://www.isciii.es/QueHacemos/Servicios/VigilanciaSaludPublicaRENAVE/EnfermedadesTransmisibles/MoMo/Documents/informesMoMo2020/MoMo_Situacion%20a%2019%20de%20julio_CNE.pdf (accessed August 2020).

14 A. Abellán García, P. Aceituno Nieto, I. Fernández Morales, D. Ramiro Fariñas, B. Castillo, and R. Pujol Rodriguez, “Una nueva estimación de población en residencias de mayores,” (2020).

15 This difference arise, among other things, because Pastor-Barriuso et al. [6] extracted deaths from the Spanish Monitor of Mortality which only registers deaths coming from 93% of Spanish population. On the contrary, this work extracted deaths from Eurostat and the National Institute of Statistics which are reporting on the 100% of population.

